# Wastewater detections of Bordetella pertussis and Mycobacterium tuberculosis nucleic acids in active disease outbreak sites in the USA

**DOI:** 10.64898/2026.04.09.26350536

**Authors:** Abigail P. Paulos, Alessandro Zulli, Dorothea Duong, Bridgette Shelden, Bradley J. White, Devin North, Alexandria B. Boehm, Marlene K. Wolfe

**Affiliations:** Gangarosa Department of Environmental Health, Rollins School of Public Health, Emory University, 1518 Clifton Road, Atlanta, Georgia 30322, United States; Department of Civil and Environmental Engineering, Stanford University, 473 Via Ortega, Stanford, California 94305, United States; Verily Life Sciences LLC, South San Francisco, California 94080, United States

## Abstract

Respiratory infections caused by bacterial pathogens like *Mycobacterium tuberculosis* and *Bordetella pertussis* have increased since the COVID-19 pandemic, yet clinical surveillance of both suffers from underreporting and delayed diagnoses. Wastewater monitoring is a valuable public health surveillance tool that can help fill gaps in clinical data yet has rarely been applied to respiratory bacterial pathogens despite evidence of bacterial shedding via excretion types that enter wastewater. In this study, we investigated the possibility for wastewater monitoring of two bacterial respiratory diseases, tuberculosis and pertussis, using two case studies of wastewater monitoring for *M. tuberculosis* and *B. pertussis*. We retrospectively measured concentrations of these pathogens in wastewater samples collected longitudinally from communities with and without known outbreaks of these diseases. We designed and validated a novel *B. pertussis*–specific assay for the NAD(P) gene; *B. pertussis* nucleic acids were detected sporadically in wastewater during an identified outbreak. We used a highly specific, established assay for *M. tuberculosis* nucleic acids, and found low concentrations of the marker in wastewater that were lag-correlated with clinical incidence rates 5 weeks later. Findings support the potential of wastewater monitoring for *M. tuberculosis* and *B. pertussis* to enable identification of communities with outbreaks of tuberculosis and pertussis and provide early warning for tuberculosis.

## Introduction

Wastewater monitoring (WM) is used globally for tracking infectious diseases in communities. First deployed at scale during the COVID-19 pandemic, WM allows insight into disease occurrence independent of clinical testing and has since expanded to include a wide range of viral targets.^1–3^ However, applications to bacterial pathogens remain limited.^4–7^

Detecting bacteria in wastewater poses unique challenges. Both pathogenic and non-pathogenic bacteria are diverse and abundant in wastewater, and polymerase chain reaction (PCR) assays used to detect nucleic acid markers for WM require genetic targets that are both sensitive and species- or subspecies-specific to target the intended pathogen. This is especially challenging for bacterial pathogens, as many bacterial species share highly similar genomes and horizontal gene transfer and mutations under selective pressure can compromise assay sensitivity and specificity over time, especially applied to environmental samples.^8,9^ Interpretation is also challenging, as evidence on human shedding in wastewater-relevant excretions (e.g., stool, urine) is limited for many non-enteric bacterial pathogens.^8^ As a result, most bacterial WM studies have focused on a small set of bacterial pathogens, namely enteric pathogens such as *Escherichia coli, Salmonella* spp., and *Shigella* spp., and select sexually transmitted infections including *Chlamydia trachomatis* and *Neisseria gonorrhoeae*.^4–7,10,11^ The use of WM for detecting respiratory bacterial pathogens is more limited. The *Mycobacterium tuberculosis* complex has been detected in wastewater in Africa, India, and Canada using qPCR ^12–14^, yet assay specificity and sensitivity remain key challenges. *Bordetella pertussis* has been detected in wastewater in China via qPCR, but required a specialized sample processing approach that may limit scalability for high-throughput monitoring programs.^15^

Here we explore the use of WM to identify infectious diseases caused by two respiratory bacteria, *Mycobacterium tuberculosis* and *Bordetella pertussis*. Although both are primarily transmitted via aerosols, the bacteria can enter wastewater via shedding in saliva, sputum, and stool.^15–19^ *M. tuberculosis* causes one of the world’s deadliest infectious diseases, tuberculosis (TB), causing an estimated 9,000 cases in the U.S and 1.25 million deaths worldwide in 2023.^20,21^ Whooping cough cases, caused by *B. pertussis*, have surged globally despite vaccination efforts.^22,23^ Antibiotic and multidrug-resistant strains of both pathogens are emerging globally, further increasing health risks.^24–27^ While *M. tuberculosis* and *B. pertussis* have been detected in wastewater, their low shedding rates may limit sensitivity, and genetic similarity to related species present unresolved challenges to specificity.^12–15^

In this study, we 1) develop and implement a novel droplet digital (dd)PCR assay specific to *B. pertussis*, 2) deploy TB and *B. pertussis* wastewater monitoring during active outbreaks, and 3) compare wastewater detections to clinical cases.

## Materials and Methods

### Pertussis assay design

We designed an assay specific to *B. pertussis* to target the NAD(P)-binding protein (1 copy/genome). The assay was designed by first identifying gene sequences unique to *B pertussis* that were not present in other *Bordetella* species. The NAD(P)-binding protein gene was identified as unique to *B. pertussis* using EDGAR3.0 geneset search by including all available *B. pertussis* genomes and excluding all other *Bordetella* species.^28^ This generated a gene set exclusive to *B. pertussis*. To further ensure this gene sequence was exclusive to *B. pertussis*, the NAD(P)-binding protein gene sequence was analyzed via National Center for Biotechnology Information (NCBI) BLAST, ensuring that no similar genes were present in other organisms potentially present in wastewater. Primer3 was used to design primers for this gene, and the best candidates from these primers were chosen based on minimum Primer3 penalty score and melting temperatures.^2^ The *in silico* specificity of this assay was assessed through an exclusionary BLAST analysis that excludes the intended assay target, allowing for the determination of any potential off target amplification.

The NAD(P) assay was tested *in vitro* against nucleic acids from a large collection of respiratory pathogens to determine specificity (Table S1). When panels were composed of whole viruses or bacteria, their nucleic acids were extracted and purified as described for the wastewater solids samples (see below) and then used neat as template in droplet digital 1-step RT-PCR assays. Assays were run in a single well using the same cycling conditions and post processing using a droplet reader as for the wastewater samples in singleplex.

### Retrospective testing of wastewater

#### Sample Collection

Stored wastewater samples collected as part of an ongoing WM program were tested for *M. tuberculosis* and *B. pertussis* nucleic acids retrospectively. For each target, specific wastewater treatment plants were selected to represent “outbreak” sites with known active outbreaks for each associated disease in the sewersheds, and “control” sites in the region without reported outbreaks in the sewersheds (Table 1).

**Table 1.** Description of wastewater treatment plant sites included in study. “Target” indicates which target was tested at the site. “Type” indicates whether site is an outbreak or control site. Population is the population served by the plant. Dates are in month/day/year format.

| Site | Target | Type | Population | Dates | Frequency |
| --- | --- | --- | --- | --- | --- |
| MV, Mill Valley, CA | *B. pertussis* | Outbreak Site | 30,000 | 1/2/2024 -<br>6/18/2024 | 3x/week |
| SJ, San Jose, CA | *B. pertussis* | Control Site | 1,500,000 | 1/18/2024 -<br>6/13/2024 | 1x/week |
| KC1, Kansas City, KS | *M. tuberculosis* | Outbreak Site | 35,000 | 7/1/2024 -<br>12/31/2024 | 1x/week |
| KC2, Kansas City, KS | *M. tuberculosis* | Outbreak Site | 90,000 | 7/1/2024 -<br>12/31/2024 | 1x/week |
| KC3, Kansas City, KS | *M. tuberculosis* | Outbreak Site | 15,000 | 7/1/2024 -<br>12/31/2024 | 1x/week |
| LW, Lawrence, KS | *M. tuberculosis* | Control Site | 80,000 | 7/1/2024 -<br>12/31/2024 | 1x/week |
| SL, Salina, KS | *M. tuberculosis* | Control Site | 47,000 | 7/1/2024 -<br>12/31/2024 | 1x/week |

For *B. pertussis*, samples from two sites in the San Francisco bay area (California, USA) were tested using the NAD(P) *B. pertussis* assay. Three samples per week (n = 72 total) were tested from Mill Valley (MV), a site with a reported outbreak during the study period (1/2/2024 - 6/18/2024, m/d/y format) that serves approximately 30,000 people. One sample per week (n = 22 total) was tested from San Jose (SJ), a control site without a reported outbreak during the study period (1/18/2024 - 6/13/2024) that serves approximately 1,500,000 people. For MV and SJ, 50 mL of settled solids were collected from the primary clarifier using sterile technique and clean bottles, as described elsewhere.^29^

For *M. tuberculosis*, samples from 5 sites in Kansas, USA were tested using a previously published assay specific to the *M. tuberculosis* species that targets the region of difference 9 (RD9).^12^ Each site had one sample per week tested during the study period, from 7/1/2024 - 12/31/2024 (n = 28 total for each site). Three sites in the Kansas City area (KC1, KC2, and KC3) were included as outbreak sites with a reported TB outbreak active during the study period. Two additional sites in Lawrence (LW) and Salina (SL), Kansas, were included as control sites; both are outside the counties identified in health alerts as part of the Kansas City area TB outbreak and located in counties that had <6 cases of active tuberculosis reported in 2024.^30,31^ For SL, 50 mL of settled solids were collected from the primary clarifier and in all other Kansas sites 24 hour composite influent samples were collected using sterile technique and clean bottles.^29^

#### Pre-analytical processing and nucleic acid extraction

All samples were maintained at 4°C and transported to the lab immediately, where they were processed within 48 hours upon arrival. Briefly, samples, including those from the primary clarifier, were settled by allowing samples to sit for 10-15 min before aspirating off the liquid fraction and transferring the solid fraction into a falcon tube. These solids were centrifuged at 24,000□×□g for 30□min at 4□°C and the supernatant was aspirated and discarded. The solids were added to DNA/RNA shield supplemented with bovine coronavirus (BCoV) as an exogenous extraction control, and homogenized. We extracted and purified nucleic acids from the resultant supernatant using Chemagic Viral DNA/RNA 300 Kit H96 (PerkinElmer, Shelton, CT) followed by inhibition removal (Zymo OneStep PCR Inhibitor Removal Kit, Irvine, CA). A separate aliquot of dewatered solids was also dried at 110□°C for 19 to 24□h to determine the solids dry weight. These methods are provided in greater detail elsewhere and in other publications.^32,33^ Separate nucleic acid aliquots were stored at -80°C for 25-61 weeks prior to retrospective testing for *B. pertussis* or *M. tuberculosis* and SARS-CoV-2 N gene, and were thawed at 4°C before PCR analysis. Samples typically underwent 1 thaw, but in some cases, samples underwent up to 2 freeze-thaws. The SARS-CoV-2 N gene was used as a control to assess storage degradation, as explained in further detail below.

#### ddPCR Methods

*B. pertussis* nucleic acids were measured in MV and SJ samples using the NAD(P) assay (Table 1). TB was measured in KC1, KC2, KC3, LW, and SL samples using the previously-published RD9 assay specific to the *M. tuberculosis* species (Table 1).^12^

**Table 1.**
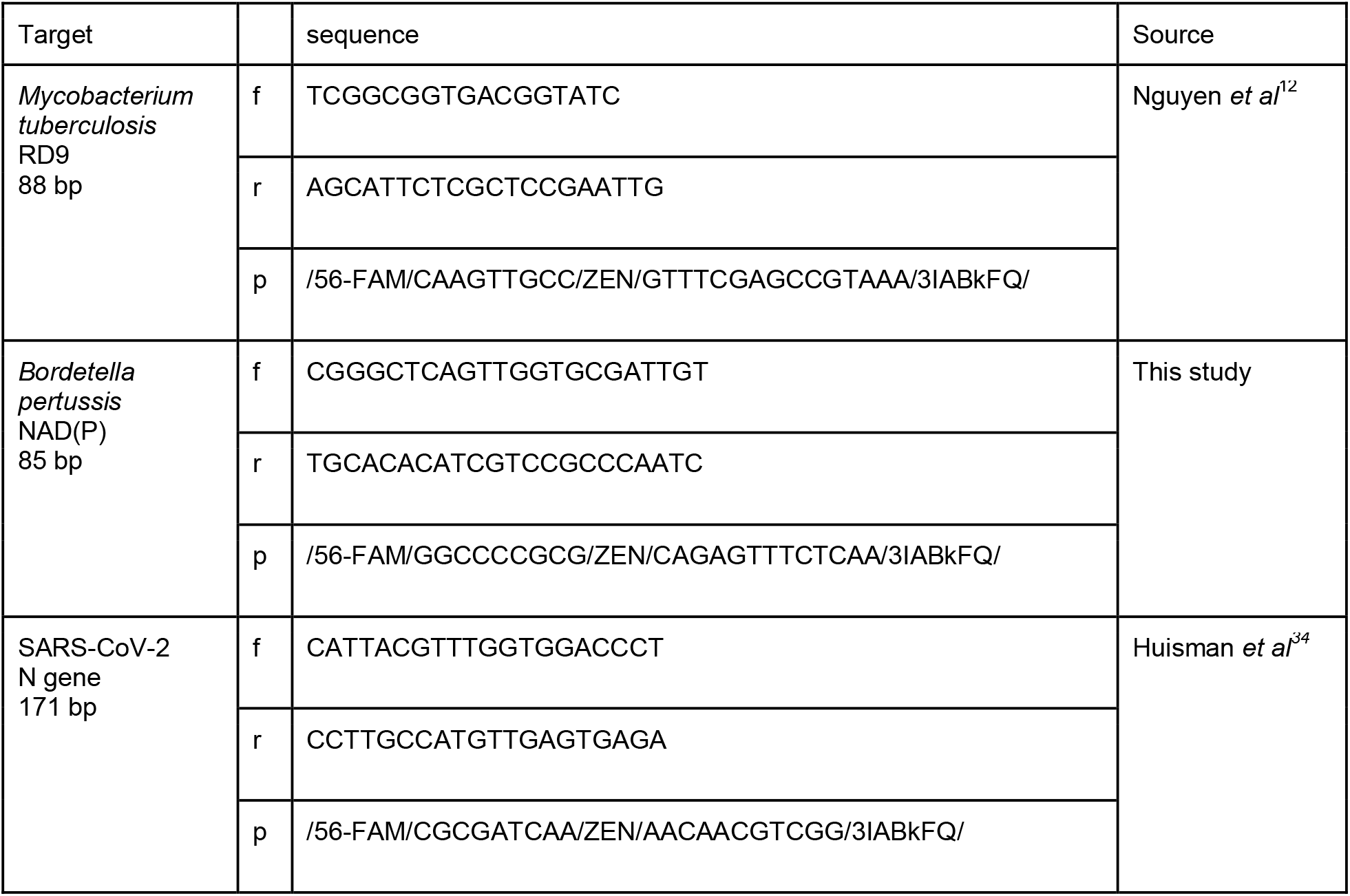
Primer and probe sequences used in this study. f, r, and p are the sequences of the forward primer, reverse primer, and probe, respectively. The probe sequence includes the fluorescent molecule and quencher.

Nucleic acids were used neat as template for RT-droplet digital PCR (RT-ddPCR). *B. pertussis* assays were run in 6 replicate wells and *M. tuberculosis* assays were run in 10 replicate wells. The NAD(P) *B. pertussis* and *M. tuberculosis* assays were each run in multiplex with the SARS-CoV-2 N gene assay using a probe-mixing approach. We used RT-ddPCR for all analyses as the goal was to ultimately multiplex the *M. tuberculosis* and *B. pertussis* assays in a panel that includes RNA-genome viruses; all assays presented here detect both DNA and RNA and results represent total nucleic acids. Fluorescent molecules used for the assays are provided in Table

1. Reaction chemistry is provided in the Supporting Information.

All targets were measured in multiplex using droplet digital one-step RT-PCR (RT-ddPCR) run on an AutoDG Automated Droplet Generator (Bio-Rad, Hercules, CA), Mastercycler Pro (Eppendorf, Enfield, CT) thermocycler, and a QX600 Droplet Reader (Bio-Rad). Only wells with over 10,000 droplets were included; all wells met this criterion. Extraction and PCR positive and negative controls were run on each 96-well plate. Positive controls consisted of quantitative genomic DNA from ATCC and custom gene blocks (Table S2). Negative controls consisted of no template controls (NTC) containing nuclease-free water.

Bovine coronavirus (BCoV) was spiked into samples and measured as an internal control. Nucleic acids were immediately assayed for BCoV and the N gene in SARS-CoV-2 without storage; these data are previously reported in the Data Descriptor by Boehm *et al*.^32^ The N gene in SARS-CoV-2 measured in samples without storage were compared to the same target as measured in stored samples to assess nucleic acid degradation.

We used QuantaSoft Analysis Pro software (Bio-Rad) to threshold droplets, and a sample had to have 3 or more positive droplets to be considered positive. Three positive droplets correspond to a concentration between 500 and 1000 copies/g, which we term the “lowest detectable concentration”. Values were converted to gene copies/dry grams using dimensional analysis. See the SI for the reaction chemistry and cycling parameters.

### Case Data

Weekly cases of whooping cough in Marin County were obtained from Marin Health & Human Services. Case dates are the earliest of symptom onset, testing date, and reporting date. Yearly case information from Santa Clara County were obtained from the California Department of Public Health (CDPH) Pertussis Surveillance Reports;^35^ case data for January 1 - April 30, 2024 were obtained from a CDPH Pertussis Snapshot.^36^ Confirmed cases require a positive laboratory test for *B. pertussis*; suspected cases with a positive PCR test but without confirmed cough are included in counts.^37^

The weekly number of newly-identified active TB cases in the Kansas City metropolitan area were obtained from the Kansas Health Alert Network.^30^ Cases are reported by the confirmation date of TB and require a diagnosis based on skin or blood tests or a chest x-ray. Total yearly cases of TB for Saline County (site of SL) and Douglas County (LW) were obtained from the Kansas Reportable Disease Statistics dashboard.^31^ Raw case numbers were converted to incident cases (cases/100,000) by dividing by the population of the Kansas City metropolitan area, 2,253,579, and multiplying by 100,000.^38^

### Statistics

N gene concentrations in samples run fresh and after storage were compared to assess potential target degradation during storage of nucleic acids. Spearman correlation tests were used to estimate the correlations between weekly clinical cases of TB or pertussis and weekly wastewater concentrations of *M. tuberculosis* and *B. pertussis*, respectively. Wastewater concentrations were aggregated to weekly data to match clinical case reporting by computing the median concentration, binary presence/absence, and for *M. tuberculosis*, the number of plants (of a maximum of 3) with a *M. tuberculosis* detection per week. Wastewater weekly values were then lagged by 0-6 weeks to determine whether wastewater concentrations precede identification of clinical cases. p values were adjusted for multiple comparisons using the Benjamini-Yekutieli method.^39^ A Kruskal-Wallis test was used to evaluate differences in *M. tuberculosis* concentrations by treatment plant. All analyses were conducted in R version 4.4.1. All data are available at https://doi.org/10.25740/gv342kn7369.

## Results

### Pertussis Assay Performance

*In silico* testing indicated that the NAD(P) assay is 100% specific to *B. pertussis. In vitro* testing revealed that neither assay (NAD(P) and RD9) cross-reacted with other common respiratory pathogens.

### QA/QC

The Environmental Microbiology Minimal Information (EMMI) guidelines were used for reporting results (Figure S1).^40^ Across all experiments, positive and negative controls had positive and negative results, respectively indicating no contamination and good assay performance. Recovery of spiked BCoV RNA were as follows: for MV, the median (IQR) recovery was 1.0 (0.83-1.2); for SJ, 2.4 (1.2-3.4); for KC1, KC2, and KC3, 0.88 (0.72-1.2); for SL and LS, 0.95 (0.80-1.3). Fractional recoveries greater than 1 likely result from measurement uncertainty associated with the amount of BCoV spiked into the samples. SARS-CoV-2 RNA concentrations were measured immediately after sample collection and again when testing archived nucleic acids to assess potential degradation during storage. Concentrations were generally higher in retrospective analyses than in initial testing, and the ratio of new to old *N* gene concentrations ranged from 0.95 - 1.81. It should be noted that bacterial nucleic acid recovery and degradation could diverge from the viral targets whose recovery and degradation are described here.

### Pertussis wastewater results

At the pertussis outbreak site MV, 9/71 samples that were analyzed for *B. pertussis* nucleic acids with the NAD(P) assay were positive (13%) (Fig 1). Concentrations ranged from 1,700 - 6,000 gc/dry g (median 2,800 gc/dry g; interquartile range (IQR) 1,800 - 2,700 gc/dry g). The peak wastewater concentration was detected on February 27, 2024. At the control site SJ, 0/22 samples tested were positive for *B. pertussis* nucleic acids.

**Figure 1.**
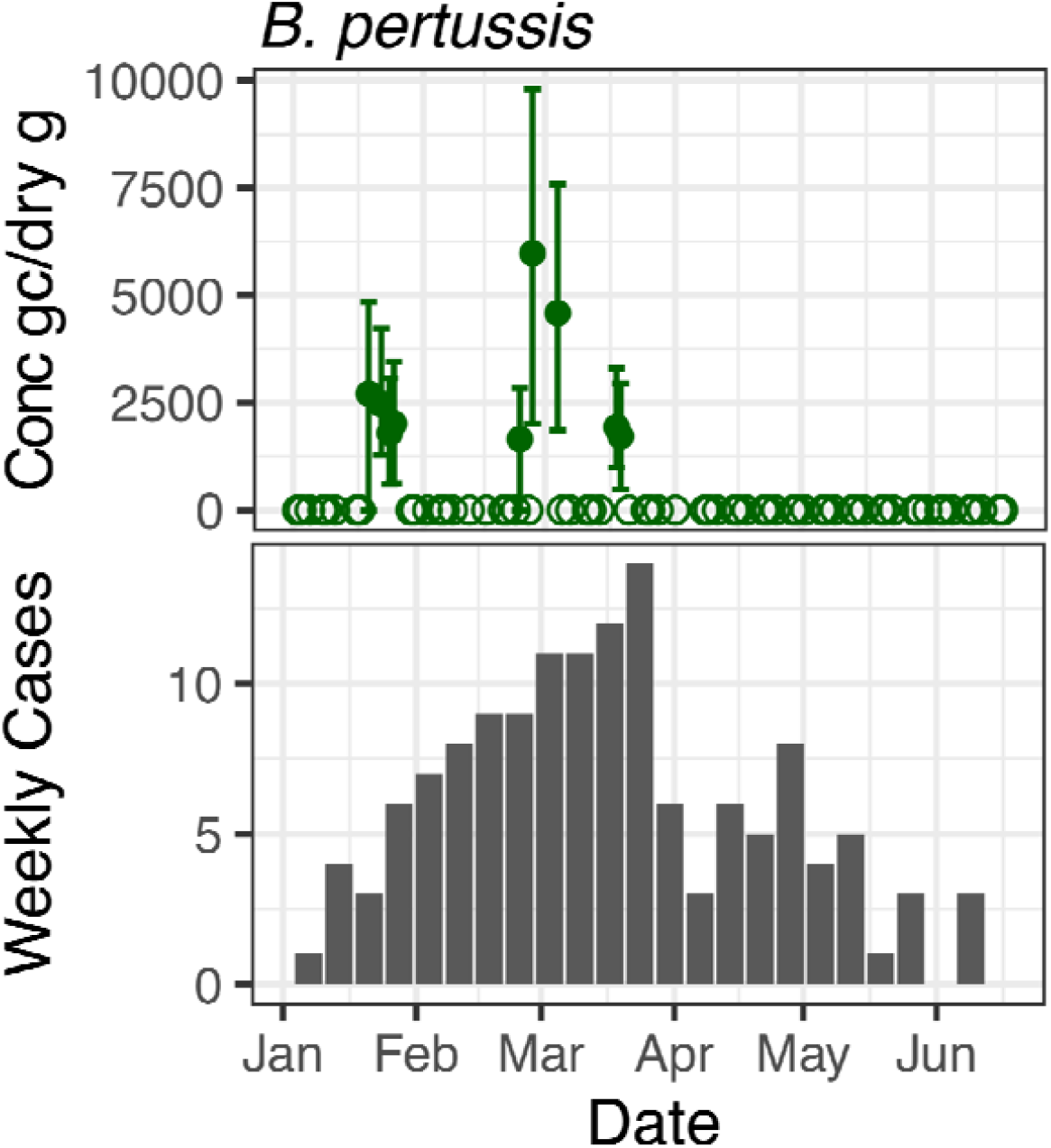
Concentrations of *B. pertussis* nucleic acids (upper) in units of copies per gram dry weight and weekly clinical cases (lower) in 2024. Open circles indicate nondetects, and filled circles indicate detections. Error bars represent 68% confidence intervals around the measured concentration.

During the time period of wastewater monitoring, between 0 and 14 cases (0 and 0.6/100,000 incident cases) per week were reported in the outbreak area, Marin County (Fig 1). The peak in case reporting was in MMWR week 13, 3/24/24 - 3/30/24. We find no correlation between clinical cases in Marin County and weekly wastewater detections (Table S3). No pertussis outbreaks were reported to the CDPH during the monitoring period in the control location, Santa Clara County; however, six cases were reported in the county between Jan 1 - April 30, 2024 and a total of 67 cases were reported in 2024.

### TB wastewater results

Throughout the study period, *M. tuberculosis* nucleic acids were detected regularly in the Kansas City outbreak sites, but not in the control sites outside of Kansas City. Overall, 17/84 (20.2%) samples from the Kansas City sites tested for *M. tuberculosis* were positive for *M. tuberculosis* nucleic acids; 7/28 (25%) at KC1, 6/28 (21.4%) at KC2, and 4/24 (14.3%) at KC3 (Fig 2). All samples from the control sites LW and SL were non-detect for *M. tuberculosis* nucleic acids. Of positive samples, the median (interquartile range) concentration was 1,640 (1,270 - 1,910) gc/dry g. Concentrations at the 3 WWTPs were not significantly different from one another (Kruskal-Wallis test, p = 0.57). *M. tuberculosis* nucleic acids were detected across the entire 6-month period of monitoring, with slightly more detections (11/42) in Jul – Sep compared to Oct – Dec (6/42), although this difference was not statistically significant (Kruskal-Wallis p = 0.14). *M. tuberculosis* nucleic acids were detected in at least one sample per week across KC1, KC2, and KC3 for 17/27 (63%) weeks of monitoring.

**Figure 2.**
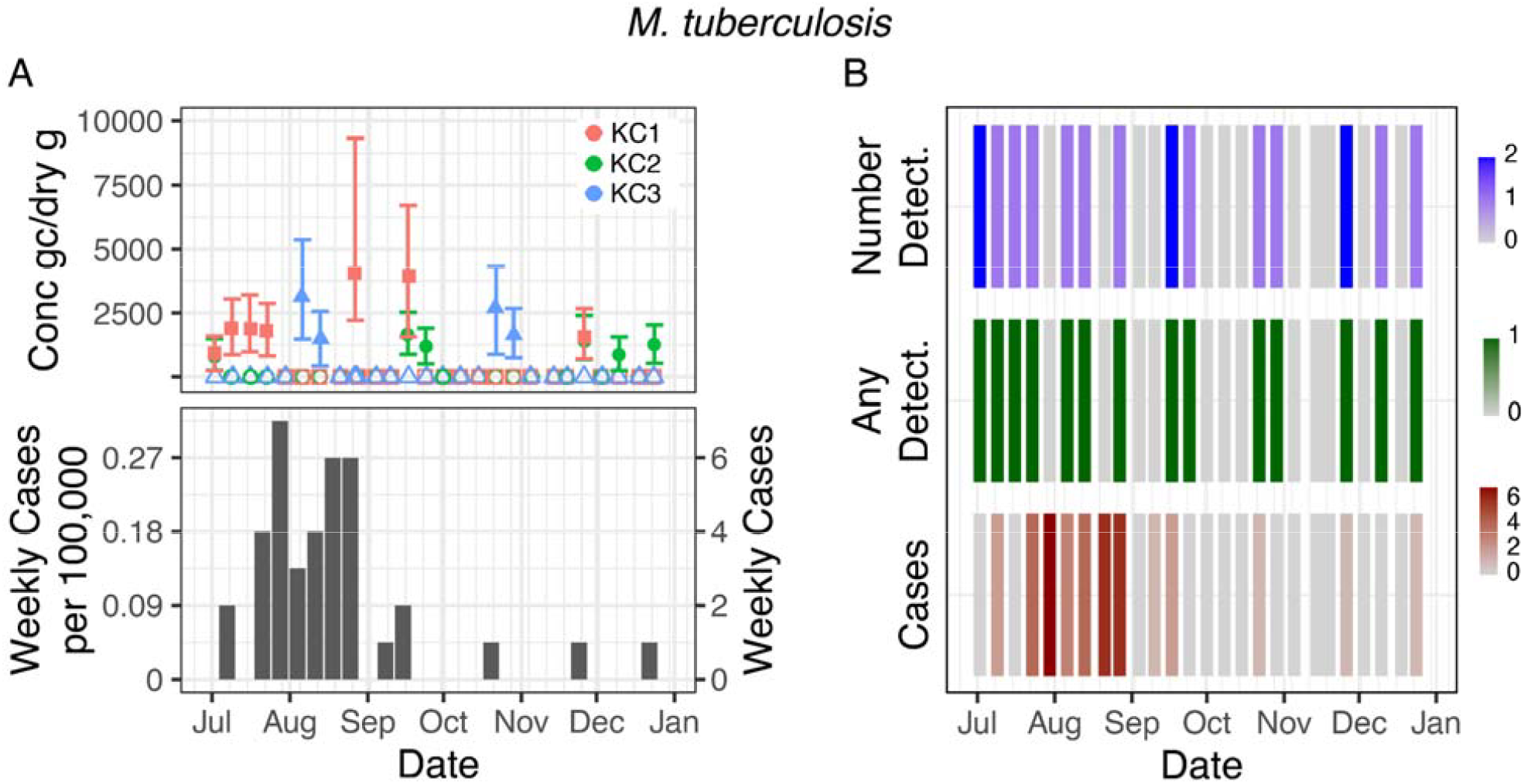
*M. tuberculosis* nucleic acid concentrations in wastewater solids (copies/g dry weight) and TB clinical cases in 2024. A) Wastewater concentrations of *M. tuberculosis* nucleic acids at the 3 treatment plants (KC1, KC2, and KC3) and weekly clinical cases of TB. Filled circles indicate detections while open circles indicate nondetects. B) Heat map of weekly wastewater detections and weekly clinical cases. “Number Detect.” indicates the number of WWTPs positive for *M. tuberculosis* nucleic acids out of a maximum of 3. “Any Detect.” indicates whether there was at least one wastewater detection across all three plants. “Cases” indicates the number of cases reported in the KC metropolitan area that week..

Fifty-one TB cases (2.3/100,000) were reported in the KC metro area during the study period. The first clinical case was reported in the week of July 9, 2024 (Fig 1). Over half of all cases (32/51) were reported in July and August. One case per month was reported in October, November, and December. In our control sites, <6 cases were reported in Douglas County (LW) and 0 in Saline County (SL) in 2024.^41^

The number of KC plants with a wastewater detection per week (of a maximum of 3) was correlated with clinical cases in the Kansas City metropolitan area 5 weeks later (n=26, r = 0.67, p = 0.0013), as was the presence of any wastewater detections at any of the 3 KC plants (n= 26, r = 0.65, p = 0.0015). No other lag periods considered (0-4 and 6 weeks) had significant associations. The median weekly wastewater concentration was not correlated with clinical cases for any lag period (Table S4 & S5).

## Discussion

In this study, *M. tuberculosis* and *B. pertussis* nucleic acids were detected in the wastewater of communities with ongoing outbreaks. While monitoring of these pathogens can provide valuable information on these diseases in the community, there were challenges with assay sensitivity and underreporting of clinical cases.

### Pertussis

Using a custom assay, we detected *B. pertussis* nucleic acids near the limit of detection at a location experiencing an outbreak of pertussis, although weekly wastewater concentrations were not correlated with reported cases in the county. However, the true case burden of *B. pertussis* is unknown; whooping cough cases are commonly underreported by 1-2 orders of magnitude, and increased clinical testing during known outbreaks means that trends in clinical cases in Marin County may not be reflective of the true case counts.^42–47^

Previous wastewater monitoring for *B. pertussis* nucleic acids found more frequent detections of *B. pertussis* nucleic acids in wastewater using a different assay, *ptx*.^15^ However, there were key differences in methodology; in the previous study, wastewater was enriched for *B. pertussis* using antibiotic exposure, a method that is intended to improve the sensitivity of detection but is not feasible in large-scale wastewater monitoring that relies on rapid, high-throughput methods for multiple pathogens at once.^15,48^

### TB

*M. tuberculosis* nucleic acid detections in wastewater during an active TB outbreak preceded clinical case reporting by 5 weeks, demonstrating the potential of wastewater monitoring to provide early warning for TB. Detections occurred sporadically across three Kansas City treatment plants, with no significant differences in concentrations or detection frequency among sewersheds. This five-week lead time aligns with prior studies documenting diagnostic delays of several weeks in the U.S.^49,50^

While most TB infections are latent, we expect that only active TB would be shed into wastewater. The disease course of TB is unique, with the majority of infections classified as latent, where a person is infected with the *M. tuberculosis* bacteria but the infection is contained and does not cause disease or spread to others. Only 5-15% of cases progress to active TB infection, and only active TB is transmissible; therefore, shedding is only expected in active cases.^12,51^

Although TB WM can fill important surveillance gaps, observed detections were near the assay limit of detection despite a relatively large outbreak for the U.S., and monitoring may be more useful in regions of the world with greater TB incidence than observed here. Even using a highly sensitive ddPCR approach with 10 replicate wells per sample, *M. tuberculosis* nucleic acid levels remained low and were just above the limit of detection. *M. tuberculosis* can be shed via feces, but the extent of fecal shedding is thought to correspond with gastrointestinal tract (GI) infection, which is only reported in 20% of active cases.^12,51^ Based on the limited detections, we suggest that the utility of TB WM in the U.S. may be limited to detection of relatively large outbreaks (tens of cases) rather than identification of individual cases.^52^ TB WM may be most valuable in monitoring trends in TB cases in high-burden regions of the world where TB cases and disease incidence are higher;^12–14^ incidence here reached a maximum of 0.3 cases/100,000, while incidence in other parts of the world such as South-East Asia and Africa can exceed 100 cases/100,000.^53^ For example, TB DNA was detected in wastewater from India^12^ and Sub-Saharan Africa.^13,14^

### Wastewater monitoring for respiratory bacterial pathogens

Temporal comparison between wastewater detections and clinical case reports revealed that *M. tuberculosis* nucleic acids were consistently detected up to five weeks before peak case reporting, consistent with known diagnostic delays for TB.^49,50^ In contrast, *B. pertussis* nucleic acids were detected intermittently before and during the peak in reported cases, with no significant association between wastewater concentrations and clinical cases. Together, these findings suggest that wastewater monitoring of bacterial pathogens could complement clinical surveillance by providing early warning of transmission or identifying circulation otherwise missed by traditional reporting but that these bacteria are found in low concentrations in wastewater solids even during active outbreaks.

These case studies highlight both the promise and ongoing challenges of wastewater monitoring for non-enteric bacterial pathogens. For *M. tuberculosis, B. pertussis*, and many other bacterial species, human shedding data are sparse, making the likelihood of wastewater detection difficult to predict.^8^ Future work should quantify bacterial shedding across wastewater-relevant excretions, including feces, urine, sputum, and saliva. Assay design also remains a critical limitation. Closely related bacterial species often share high genomic similarity, complicating the development of specific molecular targets. Further, while *M. tuberculosis* has been shown to be detectable in wastewater solids but not liquids,^12^ future studies are needed to investigate the partitioning and persistence of both *M. tuberculosis* and *B. pertussis* in varied wastewater.

In this study, we used RT-ddPCR to quantify nucleic acids of both *M. tuberculosis* and *B. pertussis*. This approach detects both RNA and DNA, which complicates interpretation of wastewater concentrations in terms of genome copies while potentially increasing sensitivity in terms of detection. Future studies quantifying the relative contribution of RNA versus DNA for these targets in wastewater would improve interpretability of measured concentrations. In addition, clinical case data are aggregated at the county or metropolitan-area level rather than the sewershed level, creating spatial mismatch between wastewater catchments and case reporting that may weaken observed relationships between wastewater concentrations and clinical incidence.

Finally, both TB and pertussis suffer from underreporting and diagnostic delays, which obscure relationships between wastewater concentrations and reported case counts. Nonetheless, WM has already proven its public health value for viral pathogens, and our findings extend that potential to two underreported bacterial diseases. We recommend that future bacterial WM efforts go beyond technical validation to include assessments of data utility, public health actionability, and integration with clinical and epidemiologic surveillance.

While wastewater monitoring for bacterial pathogens is currently limited by sparse shedding data and diagnostic uncertainties, the results here provide empirical support that even low-shedding, clinically underreported bacteria can be detected in municipal wastewater, underscoring the potential value of expanding WBE beyond viral targets.

## Data Availability

Wastewater data generated for this article as well as matched case data are available at https://doi.org/10.25740/gv342kn7369.

## Acknowledgements

This work was supported by a gift from the Sergey Brin Family Foundation to ABB. We thank the participating utilities for providing samples for this study. We thank Marin County Public Health for providing case data.

