## Supplementary Information for "Wastewater detections of Bordetella pertussis and Mycobacterium tuberculosis nucleic acids in active disease outbreak sites in the USA"

***ddPCR Reaction Chemistry and Cycling Parameters***

Briefly, 20 μl of a 22 μl reaction volume are used for ddPCR; the reaction consists of 5.5 μl template, 5.5 μl of One-Step RT-ddPCR Advanced kit for Probes, 2.2 μl reverse transcriptase, 1.1 μl dithiothreitol, and primers and probes at a final concentration of 900 nM and 250 nM, respectively. The AutoDG Automated Droplet Generator was used to generate droplets. PCR was performed using Mastercycler Pro with the following protocol: reverse transcription at 50 °C for 60 min, enzyme activation at 95 °C for 5 min, 40 cycles with 1 cycle consisting of denaturation at 95 °C for 30 s and annealing and extension at 59 °C for 30 s, enzyme deactivation at 98 °C for 10 min, and then an indefinite hold at 4 °C. The ramp rate for temperature changes was set at 2 °C/s, and the final hold at 4 °C was performed for a minimum of 30 min to allow the droplets to stabilize. Droplets were analyzed using the QX200 Droplet Reader (Bio-Rad). All liquid transfers were performed using the Agilent Bravo (Agilent Technologies).

***Assessment of nucleic acid degradation.***

SARS-CoV-2 RNA concentrations were measured immediately after sample collection and again when testing archived nucleic acids to assess potential degradation during storage. Concentrations were generally higher in retrospective analyses than in initial testing.

Table S1. Panel of organisms used for *in vitro* specificity testing

| Adenovirus 1 | Coronavirus OC43 | Parainfluenza 2 |
| --- | --- | --- |
| Adenovirus 3 | Influenza A H1N1pmd | Parainfluenza 3 |
| Adenovirus 31 | Influenza A H1 | Parainfluenza 4 |
| *B. parapertussis* | Influenza A H3 | Rhinovirus 1A |
| *B. pertussis* | Influenza B | RSV A |
| *C. pneumoniae* | *M. pneumoniae* | SARS-CoV-2 |
| Coronavirus 229E | Metapneumovirus 8 | Coxsackievirus Type A9 |
| Coronavirus HKU-1 | Parainfluenza 1 | Parechovirus Type 1 |
| Coronavirus NL63 | |  |

Table S2. Positive control materials.

| Organism | Source | Item number or sequence |
| --- | --- | --- |
| *B. pertussis* | ATCC | 9797DQ |
| *Mycobacterium tuberculosis* | ATCC | 27294D-2 |

Table S3. Correlations between median wastewater concentrations and clinical cases for B. pertussis.

| lag (weeks) | r | p | p_adj_ |
| --- | --- | --- | --- |
| 0 | 0.36 | 0.08 | 0.22 |
| 1 | 0.46 | 0.02 | 0.10 |
| 2 | 0.50 | 0.01 | 0.08 |
| 3 | 0.49 | 0.01 | 0.08 |
| 4 | 0.52 | 0.01 | 0.08 |
| 5 | 0.35 | 0.08 | 0.22 |
| 6 | 0.36 | 0.08 | 0.22 |

Table S4. TB lag analysis: cases vs weekly wastewater detections

| Number of detections per week | | | | Any detections per week | | | |
| --- | --- | --- | --- | --- | --- | --- | --- |
| Lag (weeks) | r | p | p_adj_ | Lag (weeks) | r | p | p_adj_ |
| 0 | 0.28 | 0.17 | 1.00 | 0 | 0.31 | 0.12 | 0.76 |
| 1 | -0.07 | 0.74 | 1.00 | 1 | -0.04 | 0.86 | 1.00 |
| 2 | 0.10 | 0.65 | 1.00 | 2 | 0.25 | 0.24 | 1.00 |
| 3 | 0.13 | 0.54 | 1.00 | 3 | 0.19 | 0.38 | 1.00 |
| 4 | 0.40 | 0.07 | 0.61 | 4 | 0.40 | 0.06 | 0.58 |
| **5** | **0.66** | **0.00** | **0.02** | **5** | **0.65** | **0.00** | **0.03** |
| 6 | 0.10 | 0.66 | 1.00 | 6 | 0.08 | 0.74 | 1.00 |

Table S5. Correlations between median wastewater concentrations and clinical cases for TB.

| lag (weeks) | r | p | p_adj_ |
| --- | --- | --- | --- |
| 0 | 0.07 | 0.74 | 1.00 |
| 1 | -0.14 | 0.51 | 1.00 |
| 2 | -0.33 | 0.12 | 1.00 |
| 3 | -0.10 | 0.66 | 1.00 |
| 4 | 0.16 | 0.47 | 1.00 |
| 5 | 0.31 | 0.17 | 1.00 |
| 6 | 0.10 | 0.69 | 1.00 |

| Table S6. Additional details related to EMMI guidelines: mean and standard deviation of droplets and copies per droplet per sample. | | | | | | |
| --- | --- | --- | --- | --- | --- | --- |
| Assay | samples | mean droplets | sd droplets | mean copies per droplet | sd copies per droplet | number wells |
| B. pertussis NAD(P) | Mill Valley | 104032 | 19016 | 1.57E-05 | 1.74E-05 | 3-6 |
| B. pertussis NAD(P) control sites | San Jose | 100756 | 8696 | 0.00E+00 | 0.00E+00 | 6 |
| M. tuberculosis main sites | Kansas City Metro area | 176852 | 23440 | 9.60E-06 | 1.10E-05 | 10 |
| M. tuberculosis control sites | Salina & Lawrence, KS | 97419 | 17586 | 7.10E-07 | 3.80E-06 | 6 |

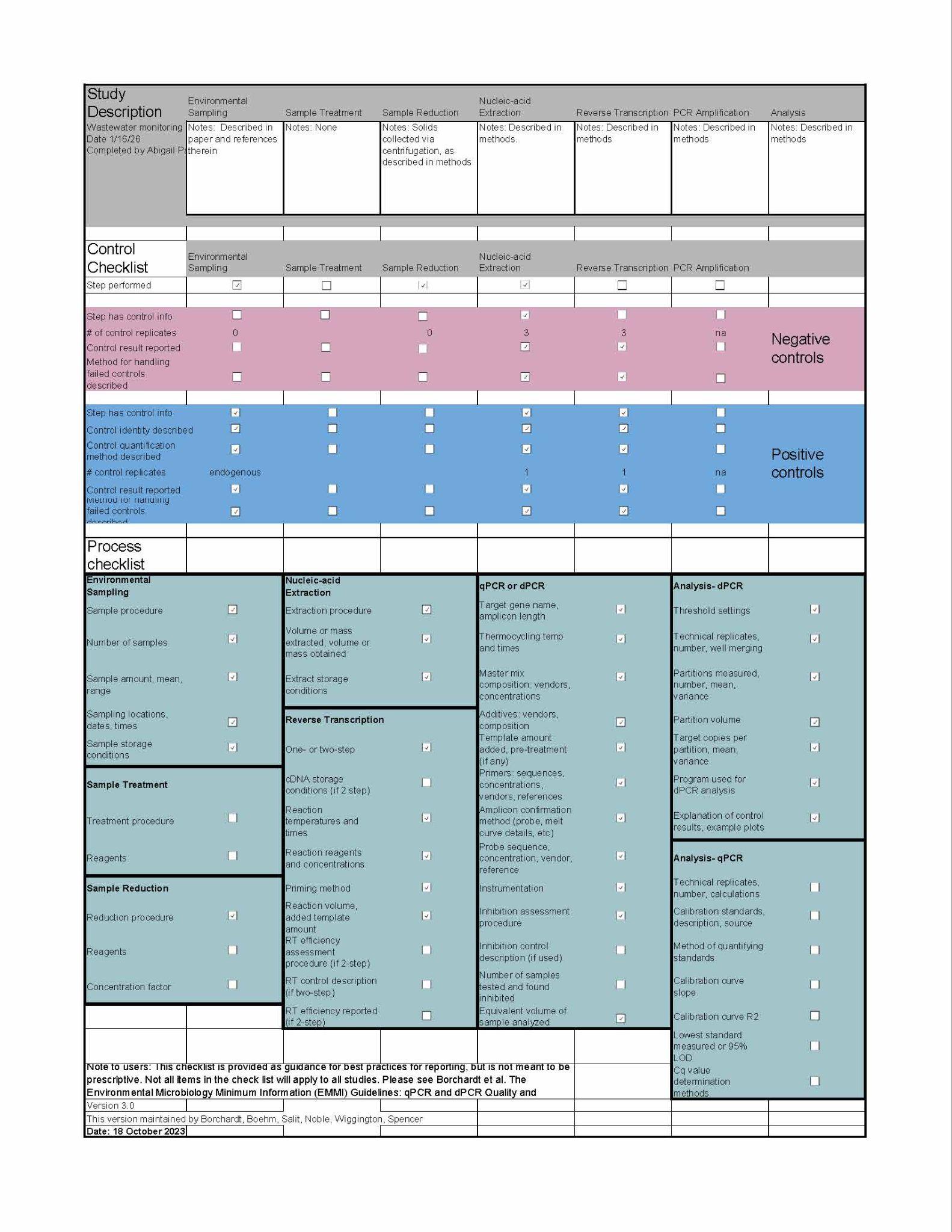

Figure S1. EMMI checklist.
